# Impact of obesity on gastroenterological diseases: a protocol for umbrella review of systematic review and meta-analysis

**DOI:** 10.1101/2020.09.17.20196386

**Authors:** Min Seo Kim, Inhyeok Lee, Yeongkeun Kwon, Sungsoo Park

**Affiliations:** Korea University College of Medicine, Seoul, Republic of Korea; Cheongsan Public Health Center, Ministry of Health and Welfare, Wando, Republic of Korea; Division of Foregut Surgery, Korea University College of Medicine, Seoul, Republic of Korea

**Keywords:** Umbrella review, meta-analysis, obesity, adiposity, body mass index, gastroenterology

## Abstract

The global prevalence of overweight and obesity among both men and women has risen as the global mean BMI has increased[1, 2]. As digestive diseases were known to have a close relation to metabolic and mechanical changes in obesity cohorts[3], several previous meta-analyses reported the association between obesity and gastroenterological diseases including cancers[4-9] and benign diseases[10-15] of the digestive system. Although some of the reported association between obesity and gastroenterological diseases might be causal, considerable heterogeneity, confounders, and bias could be included in the studies. We aim to conduct an umbrella review to evaluate the validity and strength of existing evidence for the impact of obesity on the risk and outcomes of gastroenterological (GI) diseases.

## METHODS

This investigation is currently under review for registration on the International Prospective Register of Systematic Reviews (PROSPERO). If deviations are made to the prospectively registered protocol, they will be reported in the supplementary material of final publication.

### Eligibility Criteria

#### Condition or domain being studied

Our domain of study will be multiple gastroenterological health outcomes including incidence of acute pancreatitis, cholangiocarcinoma, colorectal adenoma, diverticular diseases, gastroesophageal reflux disease (GERD), Barrett’s esophagus, gallstone, gallbladder diseases, inflammatory bowel diseases (Crohn’s disease and ulcerative colitis), non-alcoholic fatty liver disease, colon cancer, rectal cancer, hepatocellular carcinoma (HCC), esophageal cancer, gastric cancer, pancreatic cancer, and gallbladder cancer. We are also interested in the GI-specific mortality in relation to increased obesity or adiposity-related indices in general.

#### Participants/population

Participants will be classified according to standard guidelines as follows[1-2]:

- If BMI is less than 18.5, underweight.
- If BMI is 18.5 to <25, normal.
- If BMI is 25.0 to <30, overweight.
- If BMI is 30.0 to <40, obese.
- If BMI is 40 or higher, severe or morbid obese.

1. Obesity: preventing and managing the global epidemic. Report of a WHO consultation. *World Health Organization technical report series* 2000;894:i-xii, 1-253.
2. Khan, Sadiya S., et al. “Association of body mass index with lifetime risk of cardiovascular disease and compression of morbidity.” *JAMA cardiology* 3.4 (2018): 280-287.

For the analysis of the incidence of GI diseases, patients who had not been diagnosed with GI diseases prior to the initiation of the study will be included i.e. those who had not been diagnosed with the outcomes of interest (e.g. acute pancreatitis) meet the inclusion criteria.

For the analysis of the mortality, patients who had been diagnosed with GI diseases will be included in the analysis.

The incidence and prognosis of GI diseases will be analyzed separately as they involve different inclusion criteria and thereby have discrete casemixes.

#### Interventions/exposures

Exposures of interest are the increment in categorical or continuous adiposity measures (e.g. body mass index (BMI), waist circumference (WC), and waist-to-hip ratio (WHR)) as well as weight gain (kilogram) or increased visceral adipose tissue volume.

#### Comparator/control

Comparators will be the participants with normal BMI (18.5 ≤ BMI < 25) or any reference group defined per each study (e.g. lowest quantile group per BMI, WC, WHR, and other indices).

#### Types of Studies (inclusion and exclusion criteria)

Studies of the following forms will be included:

- meta-analyses of randomized controlled trials (RCT) or observational studies.
- systematic reviews with quantitative synthesis.
- studies investigated on the general population.

Studies of the following forms will be excluded:

- literature reviews with only qualitative synthesis and/or theoretical explanation.
- opinion pieces or comments.
- meta-analyses with insufficient data for analyses.
- outdated meta-analyses (more than 5 years gap from the latest meta-analysis in the identical topic)
- studies investigating underweight, weight loss, and weight loss interventions including exercise and bariatric surgery (These analyses are beyond the scope of this investigation since our research question mainly lies in how increased adiposity affects GI outcomes.)
- studies investigating the effects of adipokines on GI diseases.
- systematic reviews without meta-analyses for main outcomes.
- studies focusing on perioperative outcomes, postoperative complications, or acute consequences such as a change in biomarker and function.
- studies presenting results in prevalence rather than incidence.
- studies exclusively investigating specific populations and thus unable to extrapolate the results to general populations (e.g., child/adolescent, elderly, black, Asian, etc.)
- studies involving animal experiments or in vitro results.

### Primary Outcomes

The main outcomes will be the incidences of multiple GI diseases. Measures of the effect of obesity on multiple GI outcomes can be presented as metrics such as odds ratio (OR), relative risk (RR), hazard ratio (HR), standardized incidence ratio (SIR), and standardized mortality ratio (SMR). We will use outcome metrics identical to those reported in the original meta-analyses.

### Secondary Outcomes

The secondary outcomes will be GI-specific mortalities. When available, mendelian randomization (MR) studies will be investigated to ensure the direct causality of obesity on GI outcomes and to avoid the reverse causality whereby the presence of disease may influence adiposity. Measures of the effect of increased adiposity on GI outcomes can be presented as metrics such as OR, RR, HR, mean difference (MD), standardized mean difference (SMD), and weighted mean difference (WMD). We may convert SMD to OR if necessary.

### Search Methods for Identification of Studies

#### Electronic Database Search

We will search on PubMed, Embase, and The Cochrane Library (Cochrane database of systematic reviews) from inception to September 2020. Two researchers (MS Kim and I Lee) will independently search for systematic reviews and meta-analyses investigating the effect of increased adiposity on multiple GI diseases.

#### Pubmed

(weight* [ti] OR obes* [ti] OR adipos* [ti] OR body fat*[ti] OR overweight*[ti] OR over-weight*[ti] OR body mass*[ti] OR “body mass inde*” [ti] OR BMI[ti] OR waist circumferenc*[tiab] OR hip circumferenc*[tiab] OR “body size*” [ti] OR bariatric*[ti] OR “Body weight” [MeSH] OR “Obesity” [MeSH] OR “Adiposity” [MeSH] OR “Body fat distribution” [MeSH] OR “overweight” [MeSH] OR “body mass index” [MeSH] OR “waist circumference” [MeSH] OR “body size” [MeSH] OR “waist-hip ratio” [MeSH])

AND (oesophag*[tiab] OR esophag*[tiab] OR stomach*[tiab] OR gastr*[tiab] OR duoden*[tiab] OR jejun*[tiab] OR ileum*[tiab] OR ileal*[tiab] OR ileo*[tiab] OR cecum*[tiab] OR cecal*[tiab] OR ceco*[tiab] OR caecum*[tiab] OR caecal*[tiab] OR caeco*[tiab] OR appendix*[tiab] OR appendic*[tiab] OR vermiform*[tiab] OR vermix*[tiab] OR vermic*[tiab] OR “appendix” [MeSH] OR colon*[tiab] OR coliti*[tiab] OR pancre*[tiab] OR liver*[tiab] OR hepat*[tiab] OR bile duct* [tiab] OR bile* [tiab] OR biliar*[tiab] OR cholang*[tiab] OR biliary tract*[tiab] OR gallbladder*[tiab] OR chole*[tiab] OR rectum*[tiab] OR recta*[tiab] OR recto*[tiab] OR colorect*[tiab] OR enteri*[tiab] OR entero*[tiab] OR intestine*[tiab] OR bowel* [tiab] OR “inflammatory bowel diseas*” [tiab] OR Crohn*[tiab] OR diverticul*[tiab] OR fistul*[tiab] OR “digestive system*” [tiab] OR “esophagus” [MeSH] OR “esophageal diseases” [MeSH] OR “stomach” [MeSH] OR “stomach diseases” [MeSH] OR “duodenum” [MeSH] OR “duodenal diseases” [MeSH] OR “jejunum” [MeSH] OR “jejunal diseases” [MeSH] OR “ileum” [MeSH] OR “ileal diseases” [MeSH] OR “cecum” [MeSH] OR “cecal diseases” [MeSH] OR “colonic diseases” [MeSH] OR “colon” [MeSH] OR “pancreas” [MeSH] OR “pancreatic diseases” [MeSH] OR “liver” [MeSH] OR “liver diseases” [MeSH] OR “gallbladder” [MeSH] OR “biliary tract diseases” [MeSH] OR “biliary tract” [MeSH] OR “rectum” [MeSH] OR “rectal diseases” [MeSH] OR “gastroenteritis” [MeSH] OR “Crohn disease” [MeSH] OR “diverticulum” [MeSH] OR “fistula” [MeSH] OR “digestive system diseases” [MeSH] OR “digestive system” [MeSH])

AND (meta-analysis[ptyp] OR meta[ti] OR meta-analy*[ti] OR pooled[ti] OR mendelian*[tiab]) NOT (protocol*[ti] OR child*[ti] OR adolesc*[ti] OR young[ti] OR comment*[ti])

#### Embase

(weight*:ti,kw OR obes*:ti,kw OR adipos*:ti,kw OR ‘body fat*’:ti,kw OR overweight*:ti,kw OR ‘over-weight*’:ti,kw OR ‘Body mass inde*’:ti,kw OR bmi*:ti,kw OR ‘waist circumferenc*’:ti,kw OR ‘hip circumferenc*’:ti,kw OR ‘body size*’:ti,kw OR bariatric*:ti,kw)

AND (oesophag*:ab,ti,kw OR esophag*:ab,ti,kw OR stomach*:ab,ti,kw OR gastr*:ab,ti,kw OR duoden*:ab,ti,kw OR OR jejun*:ab,ti,kw OR ileum*:ab,ti,kw OR ileal*:ab,ti,kw OR ileo*:ab,ti,kw OR cecum*:ab,ti,kw OR cecal*:ab,ti,kw OR ceco*:ab,ti,kw OR caecum*:ab,ti,kw OR caecal*:ab,ti,kw OR caeco*:ab,ti,kw OR appendix*:ab,ti,kw OR appendic*:ab,ti,kw OR vermiform*:ab,ti,kw OR vermix*:ab,ti,kw OR vermic*ab,ti,kw OR colon*:ab,ti,kw OR coliti*:ab,ti,kw OR pancre*:ab,ti,kw OR liver*:ab,ti,kw OR hepat*:ab,ti,kw OR ‘bile duct*’:ab,ti,kw OR bile*:ab,ti,kw OR biliar*:ab,ti,kw OR cholang*:ab,ti,kw OR ‘biliary tract*’:ab,ti,kw OR gallbladder*:ab,ti,kw OR chole*:ab,ti,kw OR rectum*:ab,ti,kw OR recta*:ab,ti,kw OR recto*:ab,ti,kw OR colorect*:ab,ti,kw OR enteri*:ab,ti,kw OR entero*:ab,ti,kw OR intestine*:ab,ti,kw OR bowel*:ab,ti,kw OR ‘inflammatory bowel diseas*’:ab,ti,kw OR periton*:ab,ti,kw OR mesenter*:ab,ti,kw OR Crohn*:ab,ti,kw OR diverticul*:ab,ti,kw OR fistul*:ab,ti,kw OR ‘digestive system*’:ab,ti,kw)

AND (meta:ti OR meta-analy*:ti OR pooled:ti OR mendelian*:ti)

NOT (protocol*:ti OR child*:ti OR adolesc*:ti OR young:ti OR comment*:ti OR ‘conference abstract’:it OR ‘conference paper’:it OR ‘conference review’:it OR editorial:it OR note:it OR letter:it OR ‘short survey’:it)

#### Cochrance

(weight* OR obes* OR adipos* OR “body fat*” OR overweight* OR over-weight* OR “body mass index*” OR BMI OR bariatric*)

AND (esophag* OR oesophag* OR stomach* OR gastr* OR duoden* OR jejun* OR ileum* OR ileal* OR ileo* OR cecum* OR cecal* OR ceco* OR caecum* OR caecal* OR caeco* OR appendix* OR appendic* OR vermiform* OR vermix* OR vermic* OR colon* OR coliti* OR pancre* OR liver* OR hepat* OR bile duct* OR bile* OR biliar* OR cholang* OR biliary tract* OR gallbladder* OR chole* OR rectum* OR recta* OR recto* OR colorect* OR enteri* OR entero* OR intestine* OR bowel* OR “inflammatory bowel diseas*” OR periton* OR mesenter* OR Crohn* OR diverticul* OR fistul* OR “digestive system*”)

### Data Collection and Analysis

#### Data extraction (selection and coding)

Two researchers (MS Kim and I Lee) will independently search the existing literature using a predetermined search strategy and also manually search through reference lists of the articles found through the initial search. Studies that meet inclusion criteria will be gathered, and duplicates will be removed. Titles, abstracts, and full text of each study will be reviewed for inclusion. Ambiguous studies will be managed through discussion and agreement between two authors. If the discrepancy between the two authors persists, a third party (S Park, corresponding author) will make the decision. The study selection process will be recorded using the PRISMA flowchart (David et al, 2009).

A predefined data extraction table will be used to collect data and summarize each study. The following details will be obtained: publication year, number of included studies in a meta-analysis, number of events and cohorts, the model of effect estimation (random effects or fixed effects), heterogeneity, outcomes of interest, and evidence level (GRADE, etc).

When extracting pooled effect sizes from included studies, we prefer to use pooled effect sizes from studies that present details of all individual studies (e.g., presenting forest plot) over studies that only demonstrate summary pooled effect sizes.

#### Risk of Bias (quality) assessment

The methodological quality of included meta-analyses will be assessed using the validated AMSTAR 2 (A Measurement Tool to Assess Systematic Reviews 2) instrument. We will not examine the quality of the individual cohort studies included in the meta-analyses as this work is conducted by the authors. The certainty of evidence for each main/primary outcome will be evaluated with GRADE (Grading of Recommendations Assessment, Development and Evaluation) approach, as has been done in numerous previous umbrella reviews[16-19]. The certainty of evidence will be classified as high, moderate, low or very low. Small study effects will be assessed with Egger’s test of funnel plot asymmetry.

#### Strategy for data synthesis

We will replicate the meta-analyses in our analytic framework and re-analyze the data to uncover the non-explicit details of meta-analyses to evaluate the quality of evidence. We will re-analyze each extracted meta-analysis under both fixed and random effects model, adhere to the workflow for pairwise meta-analyses described elsewhere[20, 21]. Pooled effect sizes and 95% confidential intervals (CI) will be used for the re-analysis. We will use the same metrices as used in the original meta-analyses (RR, OR, HR, MD, SMD, and WMD). Heterogeneity between studies will be calculated as I^2^. Egger’s p-value and prediction interval will also be calculated. The results of the primary and significant outcomes (effect size and evidence level (GRADE)) will be used to construct an evidence map. Software R and its packages will be used for the analysis.

#### Subgroup and Sensitivity Analyses

Subgroup/subset analyses will be performed, when available, by these factors:

- obesity categories (overweight, obese, severely obese, etc.)
- sex
- location (e.g. Asia, Europe, etc)

The outcomes from subgroup/subset analyses are not subject to bias analyses (e.g., Egger’s test, heterogeneity test, 95% prediction interval, etc) and therefore will not be included in the evidence map/evidence level stratification.

## Data Availability

contact to

## ACKNOWLEDGEMENTS

None.

## AUTHOR STATEMENT

None

## CONFLICTS OF INTEREST

The authors declare no potential conflicts of interest.

## AUTHOR APPROVAL

All authors have been seen and approved the protocol

## Notes

### Competing Interest Statement

The authors have declared no competing interest.

